# An informatics approach to profiling patient experiences using electronic health records: constructing and clustering the burden space of individuals under 65 years of age with multiple long-term conditions

**DOI:** 10.64898/2025.11.27.25341182

**Authors:** Mozhdeh Shiranirad, Zlatko Zlatev, Roberta Chiovoloni, Emilia Holland, Jakub Dylag, Nisreen A. Alwan, Ann Berrington, Michael Boniface, Simon D. S. Fraser, Rebecca B. Hoyle

## Abstract

Living with multiple long-term conditions (MLTC) profoundly impacts patients’ lives, affecting not only their health but also their financial, emotional, and social well-being. It can impose a significant burden on people. Here we take a novel approach, exploring the lived experience of individuals with MLTC by identifying patterns of burden—spanning physical, emotional, social, and financial domains—using machine learning techniques applied to electronic health records (EHR).

We constructed a cohort of 310,990 individuals born between January 1, 1958, and December 31, 1967, all with two or more long-term conditions. Proxy indicators of burden were extracted from EHR data. Using k-means clustering, we identified subgroups of individuals with distinct burden profiles and analyzed the distribution of burden indicators within each cluster.

Several large clusters were characterized by high prevalence of one or more of pain, anxiety, and depression. Most clusters were predominantly female, with females over-represented compared to the overall burden cohort. Socioeconomic disparities were evident: clusters marked by pain had a higher proportion of individuals from the most deprived areas, while clusters characterised by stress or anxiety alone had a higher proportion of those from the least deprived areas. Certain combinations of burden indicators tended to be over-represented in the same clusters, such as pain with mobility problems, and depression with very high A&E arrivals, and separation/divorce.

This study demonstrates the utility of machine learning for uncovering nuanced, patient-centered patterns in the experience of living with MLTC. The clustering approach reveals how different burdens intersect and vary across demographic and socioeconomic lines, offering insights that could inform more tailored and equitable care strategies.

**Author summary:** Although a growing number of people are living with multiple long-term conditions (MLTCs), the nature of the burden faced by individuals and the common patterns of such person-centred burdens remain largely unknown. Previous MLTC studies have often clustered people by their long-term conditions to uncover how these conditions group together in electronic health records (EHRs). However, this approach does not capture the true complexity of MLTCs or their impact on patient experience. In this study, we identified a series of proxy burden indicators, highlighted the challenges of extracting them from EHRs, and developed data-driven methods to uncover important patterns of patient-centred burden within this large, complex space—opening new insights and a fresh research direction for understanding MLTCs. Health systems, policymakers, and clinicians stand to benefit from this study’s findings by gaining clearer insight into the expected challenges faced by different groups living with MLTCs, potentially informing more targeted support, smarter resource allocation, and better care outcomes. Researchers, in turn, benefit from a systematic methodology for clustering patient burden.

## Introduction

Over 14 million people in England live with two or more long-term health conditions (LTC), with around a third having four or more [1]. Patients with multiple long-term conditions (MLTC) account for most primary care consultations, prescriptions and hospitalisations in the UK [2].

MLTC is associated with premature mortality, prolonged hospital admissions, lower health-related quality of life, higher depression risk, and increased socioeconomic costs [3–9]. It correlates strongly with age, affecting 30% of 45-64-year-olds, 65% of 65-84-year-olds, and 82% among those aged 85+ [10, 11]. However, studies show that the number of people under 65 living with MLTC exceeds those aged 65 and over owing to the relative population sizes in these age groups [9, 12–14]. This study focuses on the experiences of this large, younger group under 65 years of age living with MLTC.

Numerous studies have been conducted using epidemiological [15–18] and data-driven [19–21] approaches to uncover patterns of MLTC. The majority focus on grouping LTCs or patients based on the number and type of LTCs or key factors influencing MLTC development [2, 22].

However, they often overlook the challenges imposed by MLTC, such as emotional, financial and logistical impacts, needs to attend frequent GP or hospital appointments or to remember to take multiple medications, potential impacts on employment or relationships [23].

We take a patient-centred perspective focusing on the “workload”, or “burden” of living with MLTC, an aspect that has not been widely studied [24], in order to inform efforts to reduce the burden of early onset MLTC.

This study is part of the MELD-B project [24]. Chiovoloni et al. [25] established the SAIL MELD-B e-cohort (SMC), using routinely gathered, anonymised, linked demographic, administrative and electronic health record (EHR) data within the Secure

Anonymised Information Linkage (SAIL) Databank [26–28]. This longitudinal e-cohort encompasses individuals resident in Wales and registered with a Welsh GP between 1st January 2000 and 31st December 2022.

To identify relevant health events, SMC participants are linked to datasets including the Welsh Longitudinal General Practice (WLGP) Dataset, the Patient Episode Database for Wales (PEDW), the Emergency Department Data Set (EDDS), and the Outpatient Database for Wales (OPDW), with data collection beginning on January 1, 1990 [25]. Holland et al [23] synthesised qualitative evidence to describe the lived experience of MLTC and conceptualise its associated burden.

In this study, we curated exploratory features within the SMC, guided by Holland et al.’s workload themes [23], to construct a “burden space” and applied unsupervised machine learning to identify burden clusters in early-onset MLTC.

We designed a customised clustering pipeline for this high-dimensional, large-scale, heterogeneous, and noisy dataset. While individual techniques are well-established, their integration forms a novel pipeline tailored to the specific challenges of our data and burden space. Its modular and adaptable structure makes it suitable for other domains with similar real-world data characteristics.

Our study offers a fresh perspective on MLTC through the lens of patient burden. The burden space and resulting clusters provide new insights into patient experience, moving beyond traditional clinical metrics to highlight the everyday impacts of MLTC.

## Methods

### 0.1 Ethics Statement

The study was conducted in accordance with the UK Policy Framework for Health and Social Care Research. Ethics approval for this study was obtained from the University of Southampton Faculty of Medicine Ethics committee (ERGO II Reference 66810). The SAIL Databank independent Information Governance Review Panel approved this study (SAIL Project: 1377). No individual informed consent was required because the dataset used in this study contained only anonymized information and no identifiable personal data.

### 0.2 Building a burden space

We construct a *burden space* using SAIL data to characterise the burden experienced by MLTC patients. The *burden cohort* includes individuals born between January 1, 1958 and December 31, 1967, ensuring *early-onset* MLTC by selecting those under 65 by the end of the SMC (December 31, 2022). This ensures cohort members lived in a similar era, with access to similar health services. All members were alive and resident in Wales on January 1, 2013, the start of the burden assessment period. To focus on individuals living with MLTC, we include only those with recorded instances of at least two LTCs (listed in Supplementary Table S2) in either the WLGP or PEDW datasets. Anxiety and depression are excluded from the LTC list but retained as proxies for psychological burden. Cohort members are followed until December 31, 2022, or until death, migration, or residency break, whichever occurs first.

Next we identify and extract burden space features for burden cohort members using an observation or look-back window from 1 January 2013 to 31 December 2022 (Figure 1b).

**Fig 1.**
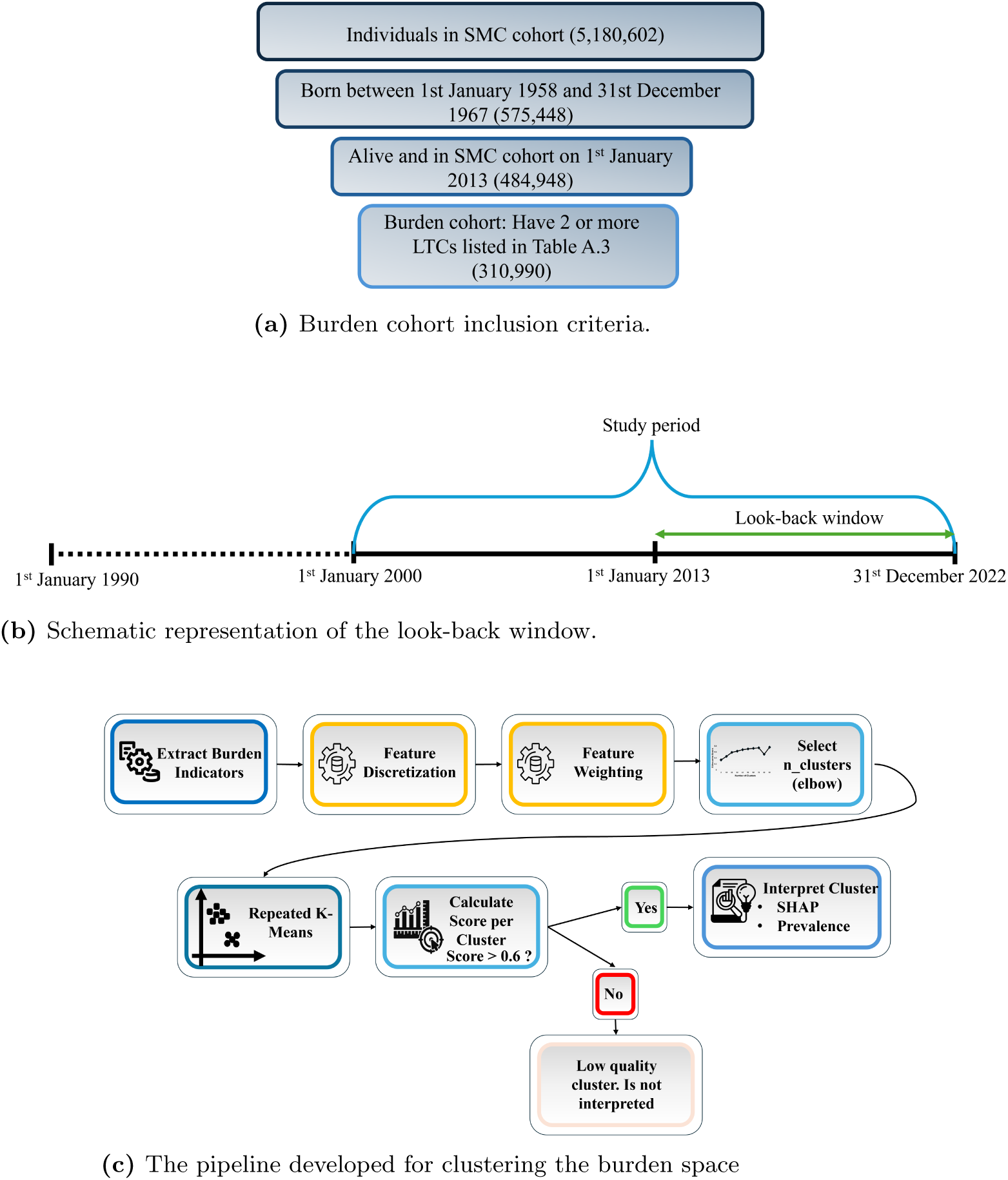
(a) The inclusion criteria for the burden cohort. (b) Look-back window used for the extraction of burden indicators. Burden indicators are extracted using the look-back window, except for the total number of LTCs and the average interval between LTC accrual, which are calculated over each individual’s full available EHR. (c) The clustering pipeline.

We created a burden space of 21 features: Anxiety, Depression, Stress, Loneliness, Pain, Mobility problems, Fatigue, Sleeping problems, Separation/divorce, Work change (changes in job or employer, redundancy etc), Unemployment, Total number of LTCs,

Number of medications, Electronic frailty index (EFI), Welsh Index of Multiple Deprivation (WIMD), Average interval between accrual of LTCs, Number of GP interactions, Number of did not attend (DNA) appointments, Number of inpatient hospital admissions, Number of outpatient hospital attendances, Number of Accident and Emergency (A&E) arrivals. These were chosen as being related to workload themes generated by Holland et al. [23] and characterisable using data available within SAIL. We consider them *proxy indicators* of burden, as SAIL EHR do not at this time routinely record patient burden directly. While we include diagnoses of depression and anxiety in our list as proxy indicators of psychological burden, we acknowledge that they are themselves long-term health conditions, and to avoid their playing a dual role in the analysis, we removed them from the list of LTCs considered (supplementary Table S2). We recognise that the presence or absence of a burden indicator record in a person’s EHR does not directly capture their experience of burden - burden indicators that are present may not always be recorded by the healthcare provider, and where burden indicators are recorded they may not always be experienced as burdensome by the patient. With these limitations acknowledged, we aim to explore what is possible within currently available EHR data, working with our exemplar set of burden indicators.

#### 0.2.1 Burden features based on clinical code lists

For features identified using clinical code list we applied the concept curation pipeline introduced by Chiovoloni et al [25], which supports the identification of records for a specific burden concept using associated clinical codes from systems such as SNOMED, Read v2, ICD-10, or OPCS-4. In SAIL the Read v2 and ICD-10 are used. Code lists for anxiety, depression and the LTC listed in supplementary Table S2 were derived from existing Read v2 code lists [25]. Remaining burden concept code lists were developed in SNOMED by the MELD-B clinical team, reviewed by at least two clinicians experienced in primary care coding [29], and converted to Read v2 for use in SAIL. These lists were used to extract binary burden indicators for anxiety, depression, stress, loneliness, mobility problems, fatigue, sleeping problems, separation/divorce, work change, and unemployment, based on the presence of any record in WLGP or PEDW during the look-back window. Supplementary Fig. S1 presents the proportion of individuals in the cohort with recorded data for each indicator.

#### 0.2.2 Engineered burden features

##### Total count of LTCs

The overall number of LTCs is determined by counting the distinct LTC diagnoses listed in supplementary Table S2 recorded in either the WLGP or PEDW dataset for each individual.

Although recovery, remission and relapse are possible for certain LTCs, determining the periods during which an LTC is active from EHR is a complex task beyond the scope of this study. Thus, we assume LTCs persist continuously from their first recorded instance and assess total LTC burden at the end of the look-back window by counting all distinct LTCs documented in the individual’s record up to that point. This same consideration means our criterion of having at least two recorded LTCs serves only as a proxy for MLTC, as we cannot be certain that the conditions co-occurred or persisted throughout the look-back period.

##### Average interval between accrual of conditions

The mean interval, in days, between consecutive first diagnoses of the distinct LTCs is calculated for each individual based on the earliest recorded instance of each LTC in either PEDW or WLGP. For interpretability, intervals are converted from days to years.

##### Welsh Index of Multiple Deprivation (WIMD)

We use the minimum Welsh Index of Multiple Deprivation (WIMD 2019) quintile over the look-back window. The index provides a score from 1 (most deprived) to 5 (least deprived) based on neighborhood-level deprivation. All cohort members had at least one WIMD record during this period.

##### Frailty

We use the electronic frailty index (eFI) developed using 36 deficit variables in primary care data [30] and validated for SAIL [31]. The eFI score reflects the proportion of identified deficits within the look-back window and categorises individuals as fit(0), mildly(1), moderately(2) or severely(3) frail [31].

##### GP interactions, DNAs, Inpatient admissions, Outpatient attendances and A&E arrivals

For GP interactions, DNAs, inpatient hospital admissions, outpatient hospital attendances, and A&E arrivals, we determine the maximum count of related records within any 12-month rolling window across the look-back period. All relevant records are considered, regardless of their association with the LTCs listed in supplementary Table S2.

For GP interactions, we count the total number of “events” recorded per patient in the WLGP Dataset, allowing a maximum of one event per day within each 12-month window ^1^. Then, the maximum count across all windows is retained.

Similarly, for outpatient attendances, we calculate the maximum number of records in the OPDW dataset over all 12-month windows. For A&E arrivals we use the maximum number of “administrative arrivals” recorded in EDDS. For inpatient hospital admissions, we count the maximum number of superspell-level admissions over all windows.

For DNAs, records were identified using a list of related clinical codes together with the concept curation pipeline, with the maximum number of DNAs per individual calculated over all 12-month windows.

##### Pain

To identify pain, we adopt a drug-frequency approach consistent with Hanlon [32] and Hafezparast [33], using a threshold of four or more prescriptions per year. Two clinician-curated medication lists are used: (1) *Pain Medications*, which includes all pain-related drugs, and (2)*Pain medications if no epilepsy diagnosis*, which are drugs used for both pain and epilepsy. Individuals are flagged as experiencing pain if they receive at least four prescriptions from either list within a 12-month rolling window. For the second list, those with an epilepsy diagnosis are excluded to ensure prescriptions reflect pain treatment.

The pain feature is set to 1 if pain is detected in any window during the look-back period, and 0 otherwise.

##### Number of medications

To capture medication burden, we count the number of ‘regularly prescribed’ medications, which better reflect sustained treatment than one-off prescriptions.

Using a medication code list based on Wilkinson et al. [34], a drug is considered ‘regularly prescribed’ within a 12-month window if it appears in at least three of its four constituent three-month intervals. All relevant prescriptions are included, regardless of their association with LTCs in supplementary Table S2.

For each individual, we compute the number of distinct regularly prescribed medications in each rolling 12-month window during the look-back period and retain the maximum.

### 0.3 Cluster Analysis

The burden space represents a complex domain, that challenges unsupervised machine learning methods due to its large-scale, high-dimensional, and noisy data, which resists analysis within existing holistic health data science frameworks. To address this, we developed a bespoke pipeline (Figure 1c), based on a repeated k-means clustering algorithm.

The burden space is a mixed-type dataset with continuous, ordinal categorical, and binary features. We discretize continuous features into ordinal categories, using the unsupervised Linde-Buzo-Gray (LBG) algorithm [35, 36], which reduces noise [37] and enhances both clustering performance and clinical interpretability. We use the *silhouette score* [38], a measure of cluster cohesion and separation, to assess cluster quality and tune hyperparameters (see supplementary S3 Appendix, S2 Appendix).

The nature of our dataset presented a challenge for cluster analysis. A clusterability check known as *separability* [39] states that an increase in the number of clusters should lead to a higher silhouette score. However this did not hold for our dataset when discretized continuous features were included. Following [40], we down-weighted these features using a simple yet effective scaling method (see supplementary S4 Appendix) to improve clusterability.

We applied the cuML GPU-enabled k-means clustering algorithm [41] with Euclidean distance metric, a scalable method suitable for use with big data and adapted for parallel architectures. To ensure reproducibility, clustering was repeated 100 times with different initial centroids, selecting the lowest objective function. This repeated k-means approach improves clustering accuracy [42]. We used k-means++ initialization [43] to improve solution quality. K-means requires the number of clusters to be specified in advance, we chose 40 based on the elbow in the silhouette score plot (supplementary Fig S3, S2 Appendix and, S6 Appendix).

Clustering stability is key to validating robustness and reproducibility, ensuring consistent outcomes from similar datasets generated from the same probabilistic source [44, 45]. We assessed stability by training two k-means models on random 85:15 dataset splits. Using an intermediate XGBoost classifier [46], the predictions from one model were compared to the other’s labels via adjuster Rand score (ARS) [47]. The resulting ARS of 0.98 indicates near-perfect agreement, confirming the clustering approach’s stability.

We interpreted clusters with per-cluster silhouette scores above 0.6, as scores above 0.5 are generally considered “reasonable” for clustering quality [48].

### 0.4 Cluster interpretation

Clusters were interpreted using the explainable AI approach SHAP [49], and a traditional frequency analysis.

#### 0.4.1 SHAP analysis

We trained an XGBoost classifier using cluster labels as targets, and computed SHAP values to identify key features characterizing each cluster. SHAP values were visualized using bar plots of average absolute SHAP values, and layered violin plots showing feature-wise distributions (Fig 2, Supplementary Fig S4). Bar plots rank features by their average contribution, while violin plots convey both magnitude and direction of effects. All calculations and visualizations were performed using the SHAP Python package [50].

**Fig 2.**
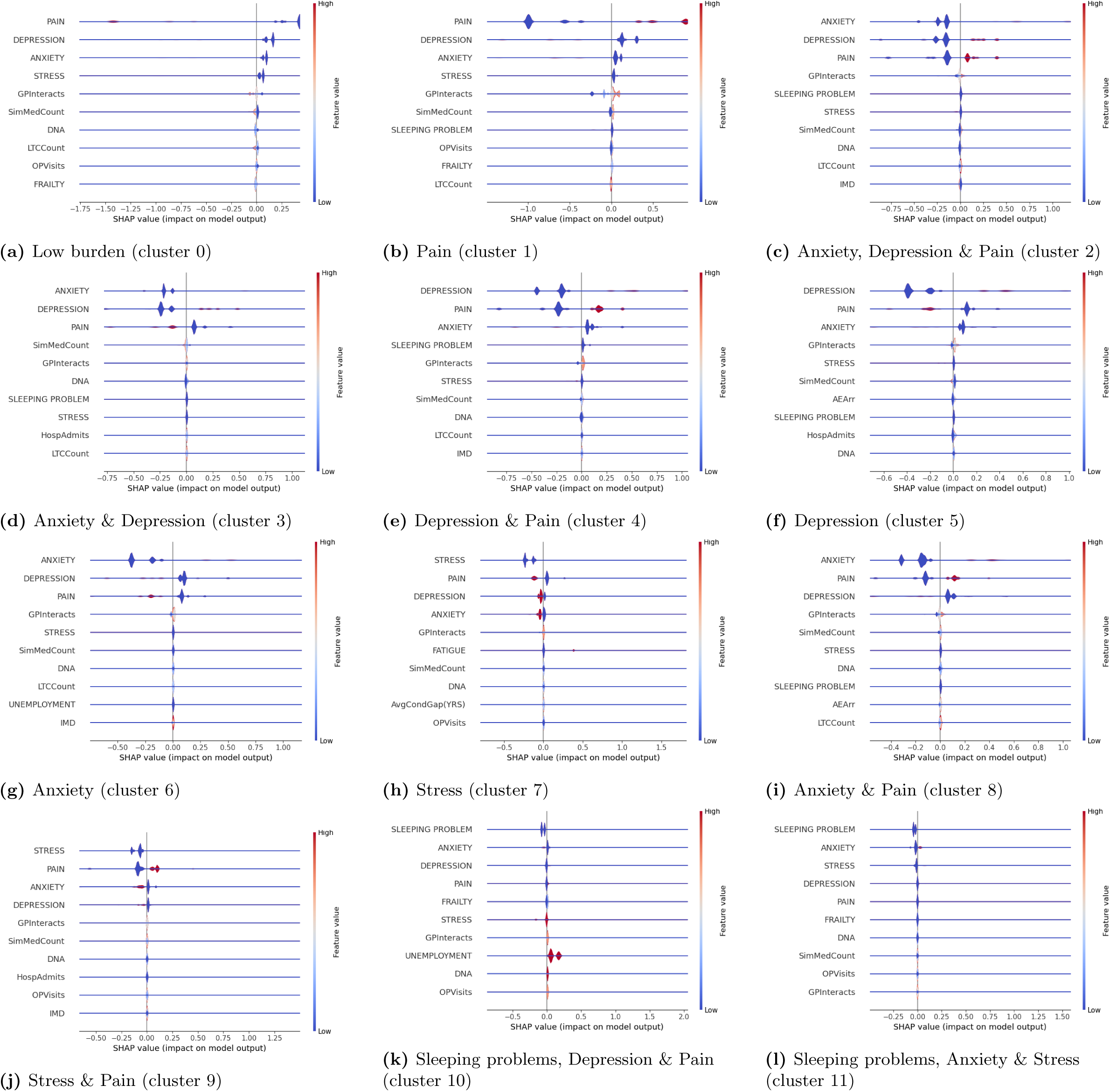
Layered violin plots of SHAP values showing the important features influencing cluster membership for all twelve identified clusters.

#### 0.4.2 Prevalence analysis

We calculated within-cluster and relative prevalence of each indicator (Supplementary section S5 Appendix). We say that an indicator is *over-represented* in a cluster if its relative prevalence is greater than one, so that it is more prevalent in the cluster than in the burden cohort as a whole.

## Results

### 0.5 Burden cohort characteristics

The burden cohort includes 310,990 members, of whom 51.6% (160,339) are female and 48.4% (150,651) are male. The majority, 88.25%, are of White ethnic background (Fig 3a), and nearly one-third reside in the most deprived areas, WIMD= 1, (Fig 3b)). The age at onset of first LTC peaks in the thirties (Fig 3c).

**Fig 3.**
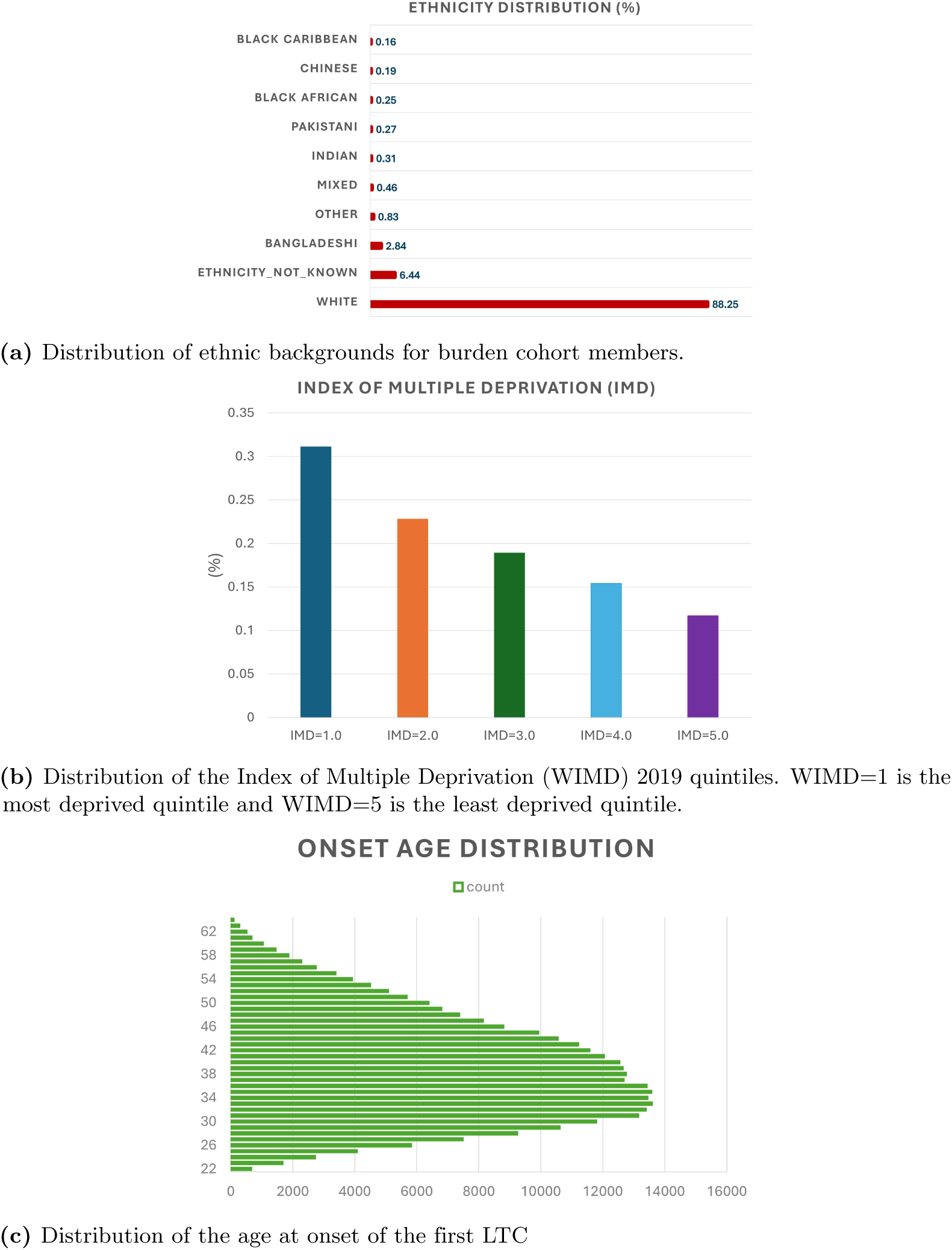
(a-c)Demographic characteristics of the burden cohort.

### 0.6 Clustering results

Using repeated k-means with an initial choice of 40 clusters, the global silhouette score was 0.47. After filtering out clusters with a per-cluster silhouette score below 0.6, 12 clusters remained (Fig 4a), resulting in a clustered population of 240,297 and an improved global silhouette score of 0.74.

**Fig 4.**
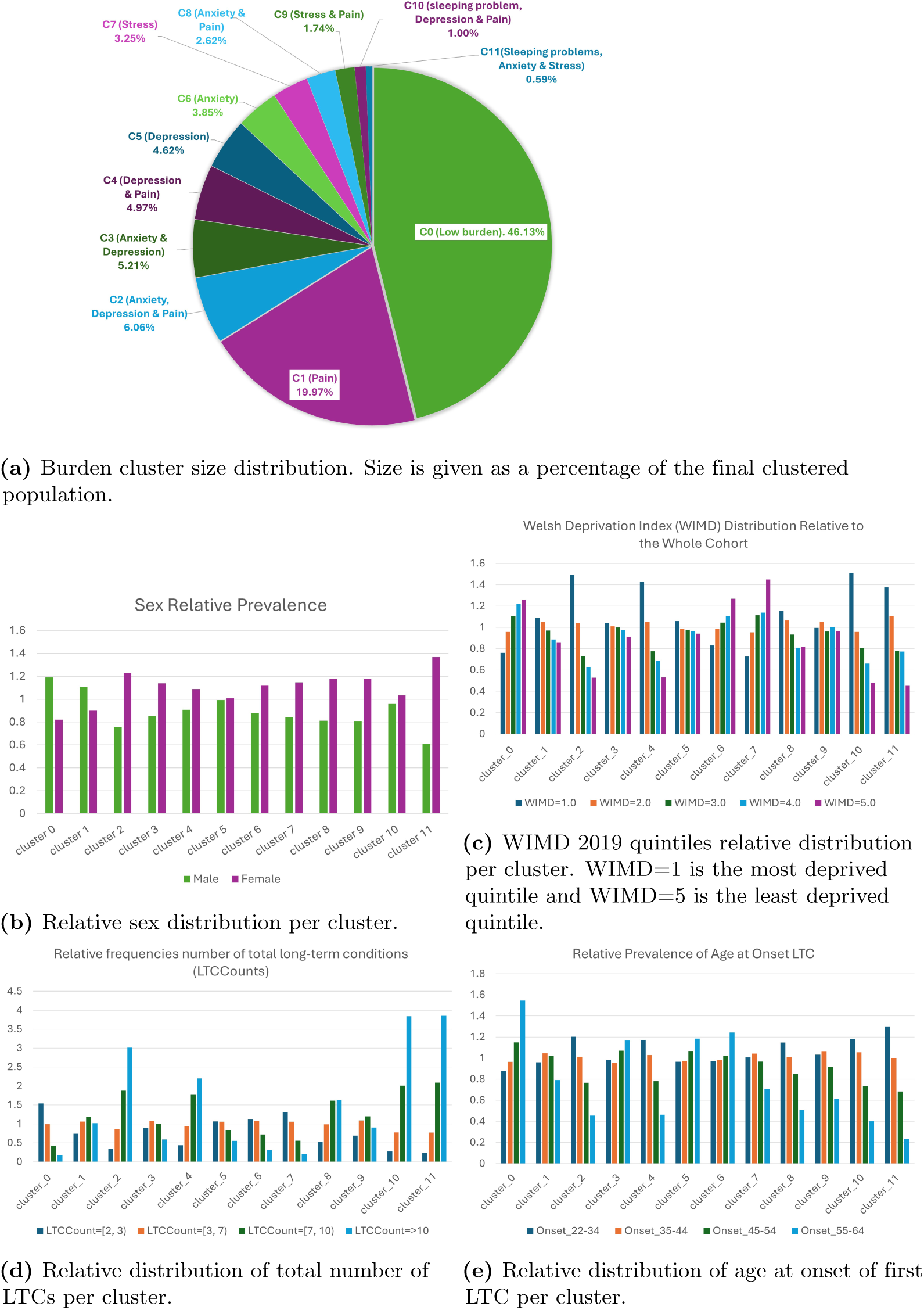
Distributions of cluster sizes, sex, WIMD 2019 quintiles, total number of LTCs and age at onset LTC per cluster. Where relative distributions are shown, they are relative to the distribution in the overall burden cohort.

All except the two largest clusters (0, “Low burden” and 1, “Pain”), are predominantly female (Fig 4b); not only do females outnumber males in these clusters, but they are also over-represented relative to their proportion in the overall burden cohort. In contrast, clusters 0 and 1 contain a higher proportion of males, who are similarly over-represented compared to the cohort as a whole.

Patients residing in the least deprived residential areas are over-represented in clusters 0, 6 and 7, whereas those from the most deprived areas are over-represented in clusters 1, 2, 3, 4, 5, 8, 10 and 11 (Fig 4c). In cluster 9, although the WIMD=2 category is slightly over-represented, the differences between WIMD groups are small.

The clusters are distinctive in terms of number of LTCs (Fig 4d): individuals with a high number of LTCs (7 or more) are notably over-represented in clusters 2, 4, 8, 10 and 11, and underrepresented in clusters 0, 5, 6, and 7, where people with lower number of LTCs are over-represented.

Older ages (above 55 years) at onset of the first LTC are over-represented in clusters 0, 3, 5, and 6, in which low or very low numbers of LTC are also over-represented (Fig 4e). We see below that cluster 0 tends to experience a lower overall burden compared to other clusters. In contrast, clusters 2, 4, 8, 9, 10, and 11 exhibit over-represented onset ages in the 22-34 age group, and are characterized by over-representation of high or very high numbers of LTCs. Except for cluster 9, these clusters are associated with higher burden, as indicated by both the number and severity of over-represented burden indicators. Cluster 7 is interesting being characterized by over-representation of both a lower number of conditions, and an age at onset of the first condition in the 35-44 age group.

### 0.7 Burden indicator SHAP and prevalence analyses of the clusters

We performed SHAP (Fig 2 and supplementary Fig S4) and prevalence analyses (Figs 5 and 6; supplementary Fig S5) of the clusters using the burden indicators.

**Fig 5.**
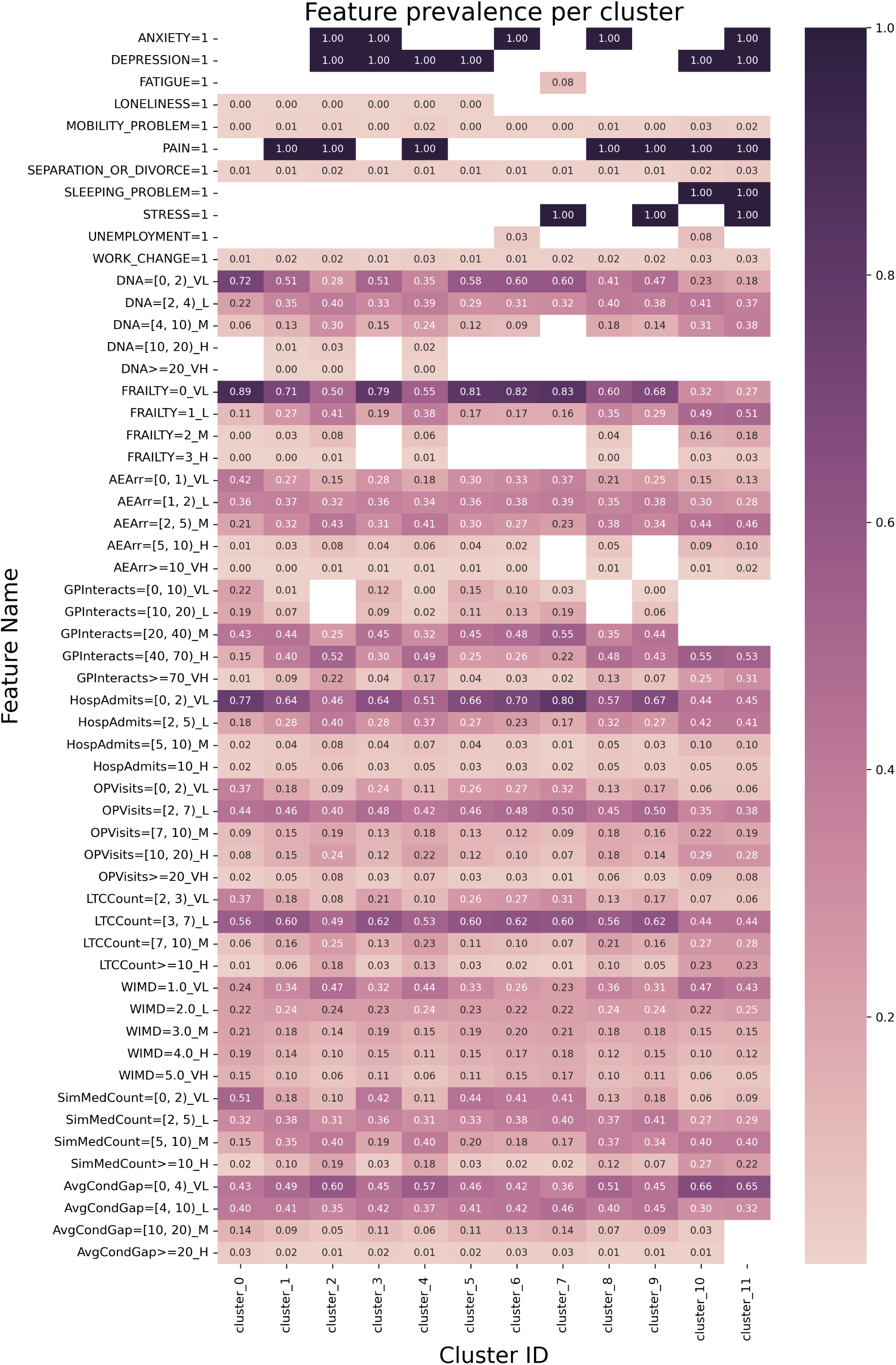
Heatmap showing burden indicator prevalence per cluster. Some cells are suppressed to mask low counts of less than 10; in some cases, extra values are masked to prevent low count identification. Key: AEArr: A&E arrivals, GPInteracts: GP interactions HospAdmits:inpatient admissions, OPVisits: outpatient attendances, LTCCount: total count of LTCs, SimMedCount: number of medications, and AvgCondGap: the average interval between the accrual of conditions.

**Fig 6.**
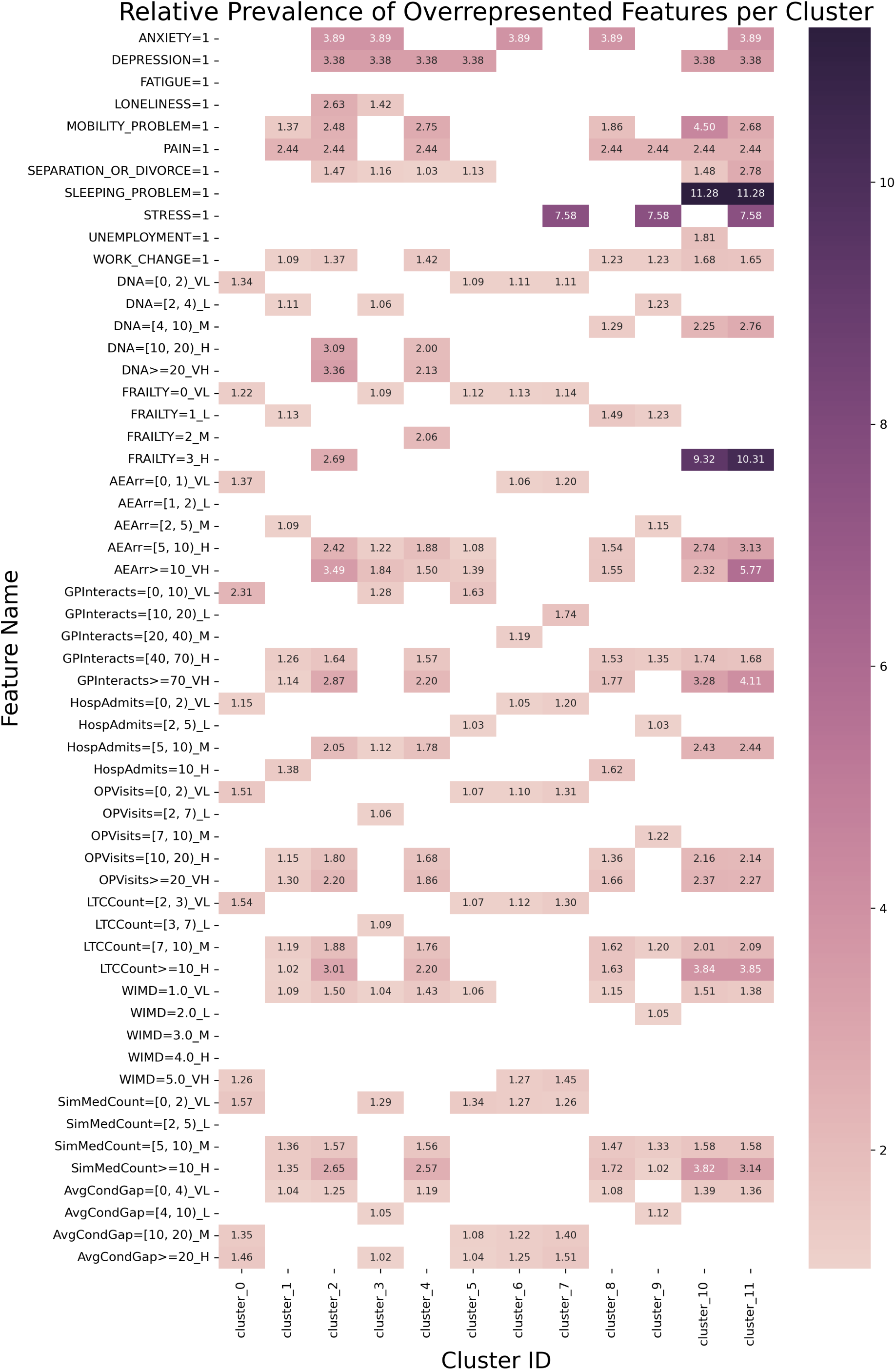
Heatmap showing the relative prevalence of burden indicator per cluster, filtered to include only values with relative prevalence *≥* 1.

Table 1 summarises the high-impact burden indicators for each cluster - down to a threshold of 0.01 for mean SHAP value - and the over-represented indicators. Clusters are named according to the binary burden indicators that are present and have SHAP values above threshold (Fig 5).

**Table 1.**
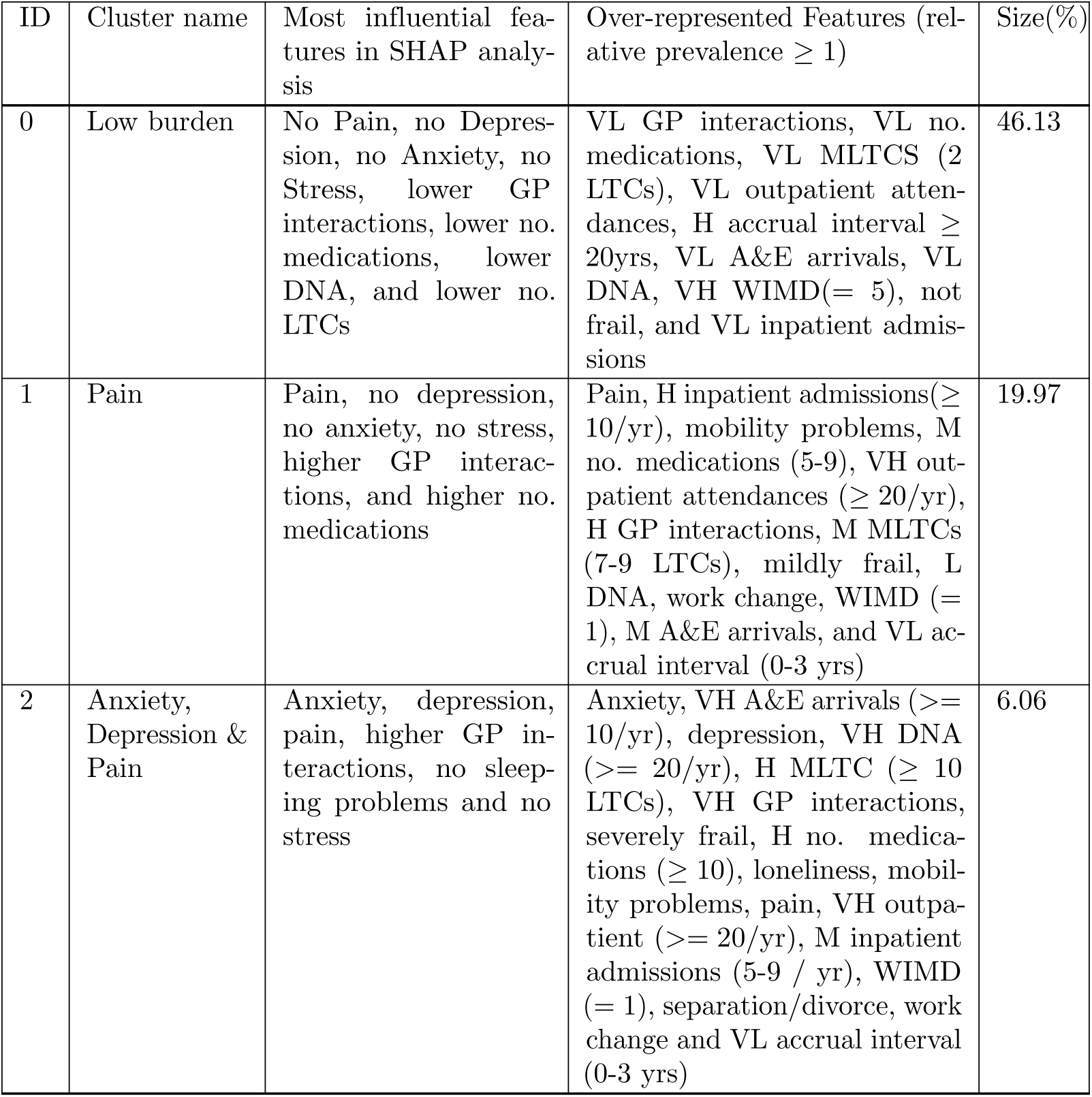
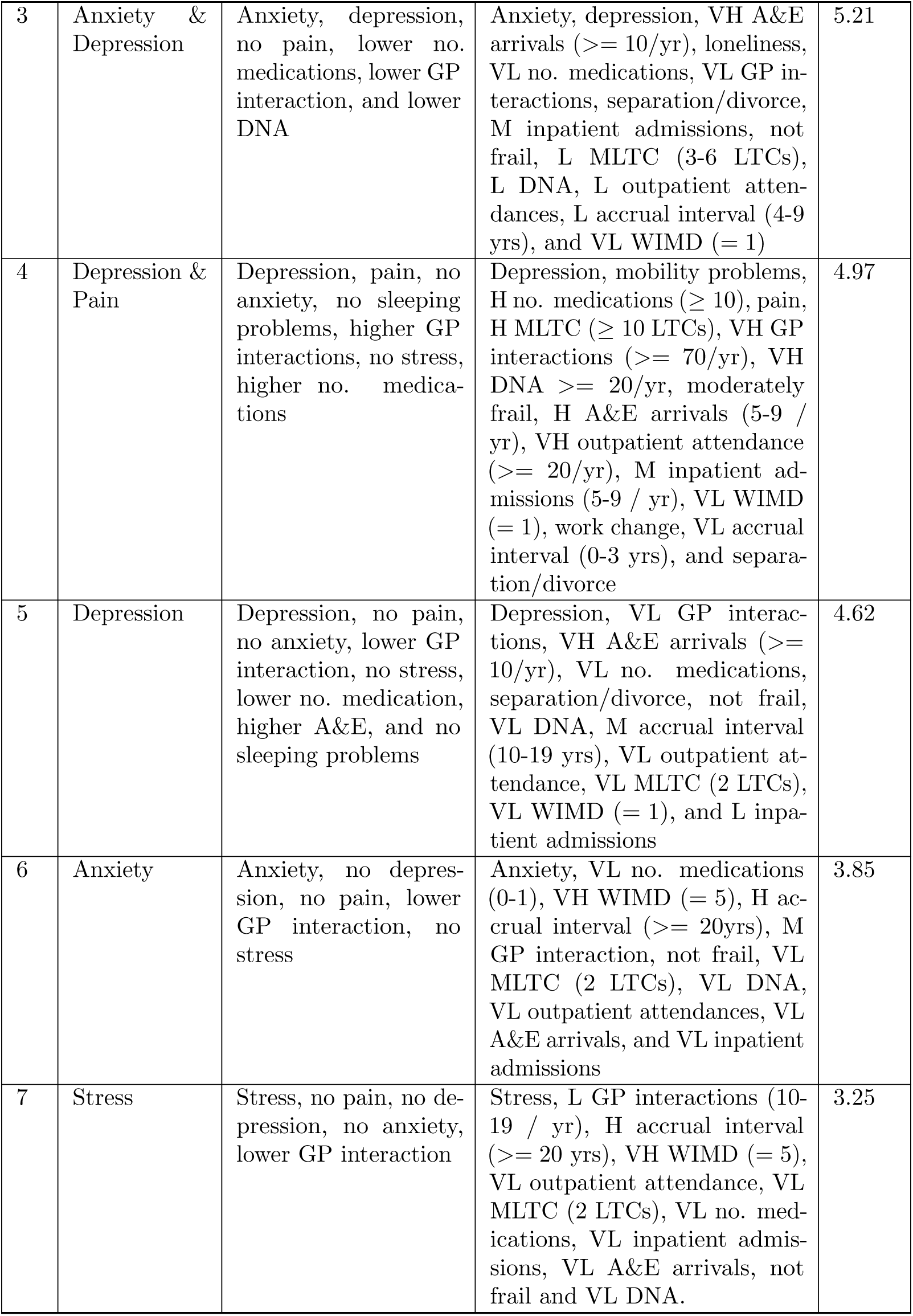
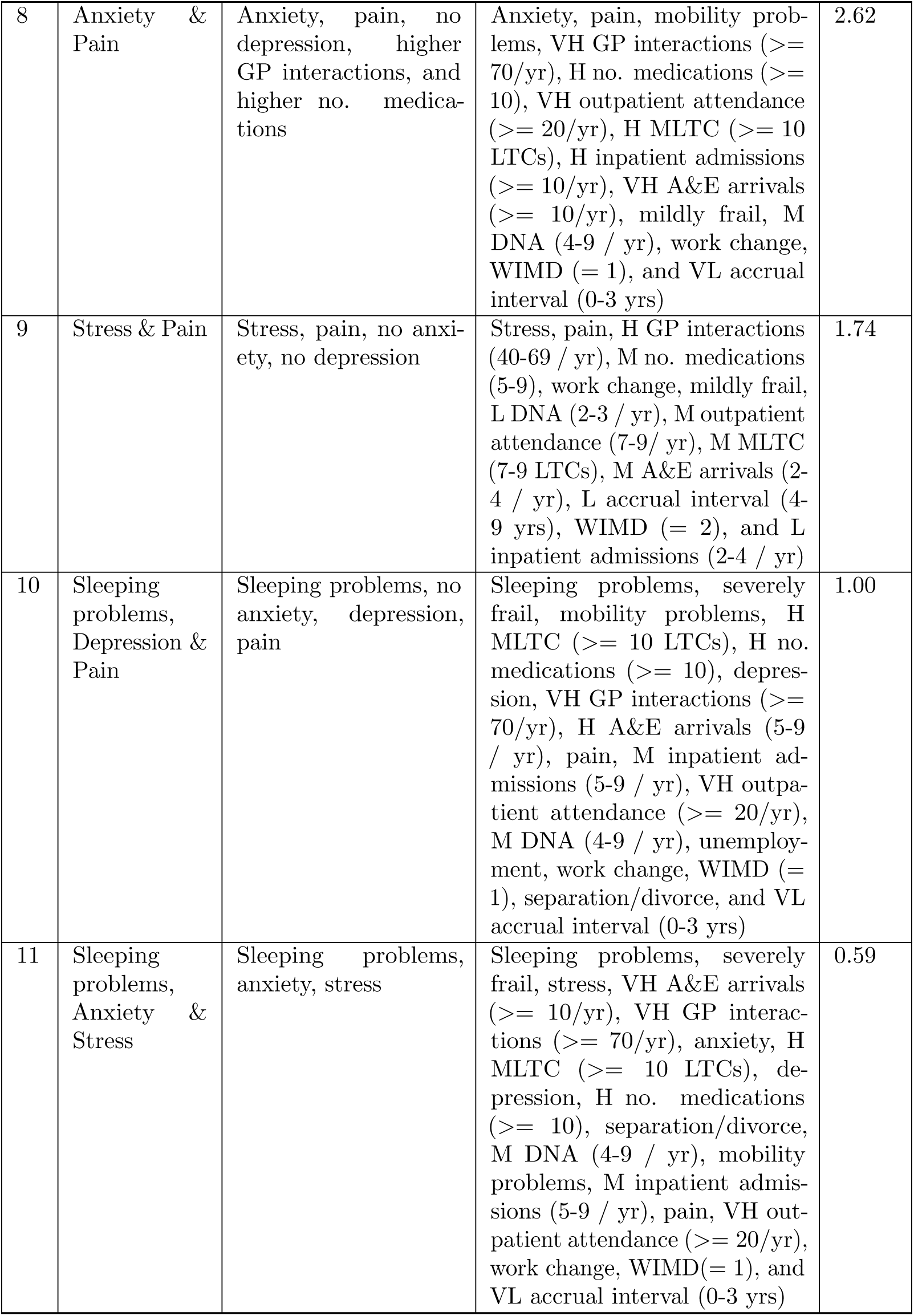
Clusters characterised by the most influential features in SHAP analysis (third column) and over-represented features in the relative prevalence analysis (fourth column), listed in order of cluster size. Influential features from SHAP analysis are listed in descending order of SHAP value, to a minimum SHAP value of 0.01. The over-represented features listed are those with relative prevalence above one. For categorical variables with multiple over-represented categories, only the category with the highest relative prevalence is reported. VL, L, M, H, and VH stand for Very Low, Low, Medium, High, and Very High, respectively. Cluster size is given as a percentage of the final clustered population.

The SHAP analysis shows that Anxiety, Depression, Pain and Stress are the driving indicators in forming most of the burden clusters.

Within the largest cluster (cluster 0, “Low burden”), patients with two LTCs, the longest time between diagnoses, and very low severity of all other burden indicators are over-represented, as are individuals from the least deprived residential areas (Fig 6). The group is predominantly male (57.65%; Fig 4b).

Within cluster 1 (“Pain”), individuals with MLTC (7–9 LTCs) and from the most deprived areas are over-represented. The other over-represented burden indicators are pain, having high levels of interaction with both primary and secondary healthcare systems, mobility problems and work change. This group is also predominantly male (53.65%).

Cluster 2 (“Anxiety, Depression & Pain”) is predominantly female (63.27%), with over-representation of those from the most deprived areas. Individuals experience severe frailty, mobility problems, extensive MLTC(*>*= 10 LTCs) with the shortest diagnosis interval, taking 10 or more medications simultaneously, and frequent healthcare use -especially GP, outpatient and A& E services. Three of the core indicators — anxiety, depression, and pain- are prevalent. In addition, the cluster exhibits over-representation of separation/divorce, loneliness and work changes.

In cluster 3 (“Anxiety & Depression”), is marked by over-representation of anxiety, depression, loneliness, separation/divorce, and frequent A&E admissions (*≥* 10/yr).

Females (58.71%), and those with MLTC (3–6 conditions) with moderate accrual interval (4-9 years) and those residing in the most deprived areas are over-represented. Despite very low GP interaction, secondary care use -especially A&E arrivals (*≥* 10/yr)-is over-represented. Medication burden is minimal, with very low number of medications over-represented.

Cluster 4 (“Depression & Pain”) shares similarities with cluster 1 (“Pain”) in over-representation of high healthcare interaction, work change and mobility problems. However, it is distinguished by high prevalence of depression, greater proportion of females (56.05%), and over-representation of the highest counts of LTCs and medications (10 or more), moderate frailty, and highest number of DNA (20 or more), likely linked to intensive GP interaction. Separation/divorce is also notably over-represented.

Cluster 5 (“Depression”) is predominantly female (51.94%), and individuals with 2 LTCs with more than 10 years accrual time gap, those from the most deprived areas and those experiencing depression, separation/divorce and very high number of A&E arrivals (more than 10 per year) are over-represented. While these burdens are notable, severity levels across other indicators are low, making this one of the least burdened clusters.

In cluster 6 (“Anxiety”), similar to cluster 5, females (57.54%), those with 2 LTCs with highest accrual time gap (more than 20 years), and those from the least deprived areas are over-represented. All cluster members have records of anxiety and moderate interaction with GP (20-39 per year) is over-represented. The overall burden profile is moderate as 9 indicators are over-represented at low or medium severity which makes it one of the least burdened groups .

Cluster 7 (“Stress”) is similar to clusters 5 and 6, with over-representation of females (59.11%), individuals with 2 LTCs, and the highest accrual time gap. In contrast, those from the least deprived areas are over-represented. Stress is the most prevalent burden. This is another least burdened cluster. This was the only cluster where fatigue played a slight role in its formation based on SHAP analysis, but it wasn’t over-represented as its relative prevalence remained slightly below one.

In cluster 8 (“Anxiety & Pain”), anxiety is the most prevalent indicator. It is similar to cluster 4 (“Depression & Pain”) in sex distribution (60.69% female) and most healthcare-related burdens. However, cluster 4 is marked by depression, separation/divorce, and higher levels of frailty, and DNAs indicating a greater personal life burden.

Cluster 9 (“Stress & Pain”) includes over-representation of females (60.81%), individuals with 7–9 LTCs, and work change. In contrast to previous pain-related clusters, individuals from less deprived areas (WIMD=2) and lower interaction with the secondary health system are over-represented.

In cluster 10 (“Sleeping problems, Depression & Pain”) includes over-representation of females (53.30%), individuals with more than 10 LTCs, severe frailty, those from the most deprived areas, and those experiencing sleeping problems, work change, unemployment, separation/divorce, and mobility problems. Compared to cluster 4 (“Depression & Pain”), individuals in this cluster are more frail. This is also the only cluster where unemployment is over-represented.

Cluster 11 (“Sleeping problems, Anxiety & Stress”) is predominantly female (70.51%). Those with 10 or more LTCs, from the most deprived areas, and the highest interaction with both primary and secondary healthcare systems are over-represented. In many aspects, it is similar to cluster 10 but includes two additional burdens -anxiety and stress-and higher A&E arrivals. This cluster does not show over-representation of unemployment. Notably, although pain and depression did not contribute to cluster formation, they are among the most prevalent indicators. Thus, all four core indicators are prevalent in this cluster, each with a prevalence of 1.

The “Low burden”, “Anxiety” and “Stress” clusters (0, 6 and 7) broadly reflect lower recorded burden, characterized by relatively low healthcare interactions, a greater likelihood of having only 2 LTCs, and very high WIMD.

The “Anxiety, Depression & Pain”, “Depression & Pain”, “Anxiety & Pain”, “Sleeping problems, Depression & Pain” and “Sleeping Problems, Anxiety & Stress” clusters (2, 4, 8, 10 and 11) show over-representation of binary burdens or severe levels of categorical ones for at least 11 of the 21 burdens, and may reasonably be described as higher-burden clusters.

### 0.8 Long-term conditions and cluster membership

We examined patterns linking LTCs to cluster membership, calculating the relative prevalence of diagnosed conditions within each cluster, focusing on the top 20 most common LTCs in the burden cohort (Fig S6), namely hypertension, drug and alcohol misuse, osteoarthritis, asthma, atopic eczema, neuropathic pain, coronary heart disease, arrhythmia, type 2 diabetes, deafness, chronic obstructive pulmonary disease, irritable bowel syndrome, hypothyroidism, chronic fatigue syndrome, diverticular disease, gout, anaemia, psoriasis, peripheral vascular disease and skin cancer. Additionally, we identified the most highly over-represented LTCs among all the LTCs in supplementary Table S2 for each cluster. Results are summarized in Table 2.

**Table 2.** Over-represented LTCs in each cluster. Column 3 shows the over-represented LTCs among the 20 most common LTCs in the burden cohort, while column 4 shows the over-represented LTCs among all LTC in supplementary Table S2. Those with relative prevalence *>* 1, up to the first five most highly over-represented LTCs are shown in each case.

| ID | Cluster name | Over-represented LTCs |  |
| --- | --- | --- | --- |
|  |  | Among 20 most common | Among all |
| 0 | Low burden | Skin cancer | Prostate cancer, Skin cancer |
| 1 | Pain | Gout, Osteoarthritis, Neuropathic pain, Peripheral vascular disease (PVD), Type 2 diabetes | Motor neurone disease, Ankylosing spondylitis, Psoriatic arthritis, Rheumatoid arthritis, Primary lung cancer |
| 2 | Anxiety, Depression & Pain | Chronic fatigue syndrome, Chronic obstructive pulmonary disease (COPD), Anaemia, Osteoarthritis, Arrhythmia | Personality disorders, Post-traumatic stress disorder (PTSD), Pancreatic disease, Fibromyalgia, Self harm |
| 3 | Anxiety & Depression | Chronic fatigue syndrome, Drug alcohol misuse, Arrhythmia, COPD, Hypothyroidism | Chronic fatigue syndrome, Obsessive-compulsive disorder (OCD), PTSD, Autism and Attention deficit hyperactivity disorder (ADHD), Eating disorders |
| 4 | Depression & Pain | COPD, PVD, Anaemia, Type 2 diabetes, Osteoarthritis | Dementia Alzheimer’s, Pancreatic disease, Multiple sclerosis, Paralysis, Chronic pain |
| 5 | Depression | Drug alcohol misuse, COPD, Hypothyroidism, Type 2 diabetes | Bipolar disorder, Dementia Alzheimer’s, Self harm, Schizophrenia, Eating disorders |

| Continuation of Table 2 |  |  |  |
| --- | --- | --- | --- |
| ID | Cluster name | Over-represented LTCs |  |
|  |  | Among 20 most common | Among all |
| 6 | Anxiety | Arrhythmia, Inflammatory bowel disease (IBS), Skin cancer | OCD, PTSD, Autism and ADHD, Arrhythmia, Parkinson's |
| 7 | Stress | Atopic eczema, IBS, Skin cancer, Deafness, Psoriasis | Long Covid, Polycystic ovary syndrome (PCOS), Endometriosis, Atopic eczema, IBS |
| 8 | Anxiety & Pain | Arrhythmia, Neuropathic pain, Osteoarthritis, IBS, Anaemia | OCD, PTSD, Ehlers-Danlos syndrome, Autism and ADHD, Plasma cell cancer |
| 9 | Stress & Pain | Neuropathic pain, Osteoarthritis, Gout, IBS, Atopic eczema | Psoriasis arthritis, Long Covid, Systemic lupus erythematosus, Chronic pain, Neuropathic pain |
| 10 | Sleeping problems, Depression & Pain | COPD, PVD, Anaemia, Type 2 diabetes, Osteoarthritis | Ehlers-Danlos syndrome, Fibromyalgia, Parkinson's, Chronic pain, Dialysis |
| 11 | Sleeping problems, Anxiety & Stress | Chronic fatigue syndrome, IBS, COPD, Anaemia, Arrhythmia | PTSD, Personality disorders, Eating disorders, Long Covid, Bipolar disorder |

Skin cancer is over-represented in the lower-burden clusters 0, 6 and 7 (Table 2, first column), which may indicate that it is associated with lower burden on average compared to the average across all LTCs considered. Along with the other over-represented condition listed for the “Low burden” cluster, namely prostate cancer, skin cancer is typically addressed quickly by the healthcare system, plausibly explaining their appearance in the cluster. Post-traumatic stress disorder appears as a common over-represented condition across all clusters where anxiety is an important burden; chronic obstructive pulmonary disease is over-represented across those where depression is prevalent; inflammatory bowel disease and long COVID across all those with stress, and chronic fatigue syndrome across those with both anxiety and depression. Among the higher-burden clusters (2, 4, 8, 10, and 11), anaemia is a shared over-represented condition among the 20 most common.

In clusters where pain is important (1, 2, 4, 8, 9, and 10), osteoarthritis is a shared over-represented condition among the 20 most common. However, these clusters differ in their most over-represented LTCs among all LTCs, which contributes to distinct burden profiles within each group.

## Discussion

Our analysis identified 12 clusters with distinct burden profiles. The SHAP analysis reveals these clusters are formed around pain, anxiety, depression, and stress, indicating that mental health, emotions and symptoms play a significant role in defining and characterizing them.

Those living in more deprived areas are over-represented in all clusters except 0, 6, and 7 (“Low burden”, “Anxiety”, and “Stress”). Among these, clusters marked by pain (1, 2, 4, 8, 9, 10 and 11) show a greater likelihood of more intensive use of the healthcare system and of having seven or more LTCs with less than 4 years gap between their diagnosis. This highlights the connection between socioeconomic deprivation and ill-health, and underscores the need for targeted support from the health system to mitigate the impacts of socioeconomic inequalities on quality of life.

Conversely, those living in the least deprived areas are over-represented in clusters 0, 6, and 7 (“Low burden”, “Anxiety”, and “Stress”) that feature greater likelihood of low numbers of conditions with more than 20 years gap between their diagnosis, and less intensive use of the public healthcare system. This perhaps suggests that higher socioeconomic status and better health are likely to be protective of each other. In addition, we speculate that recording symptom codes such as stress and fatigue may be more likely when there is no clear diagnosis, and that individuals residing in the least deprived areas may make more use of private healthcare.

Clusters 3 (“Anxiety & Depression”) and 5 (“Depression”) are centred around psychological burdens without pain, fewer than seven LTCs, and very high A&E utilization. These clusters exhibit minimal variation across WIMD levels, while the highest rate of A&E arrivals, and at the same time the lowest rate of GP interactions, are over-represented. This pattern suggests that depression may be linked to a distinct mode of healthcare utilization irrespective of socioeconomic circumstances.

Pain has been identified as a key component of patient workload by Holland et al. [23]. Three of the pain-related clusters-“Pain”, “Depression & Pain”, and “Sleeping problems, Depression & Pain” (1, 4, and 10)- show notable differences that may offer valuable insights for future research. Individuals in the latter two clusters are more likely to be female and have higher numbers of conditions, alongside additional burden records. Frailty severity, increases progressively: mild is overrepresented in “Pain”, moderate in ”Depression & Pain”, and severe in ”Sleeping problems, Depression & Pain”.

In contrast, comparing “Pain” and “Anxiety & Pain”, individuals in the latter are also more likely to be female and have more conditions, but frailty level remains unchanged. Additionally, individuals in “Anxiety & Pain” are less frail than those in “Depression & Pain”, where missed appointments (DNA) and separation or divorce are over-represented. On the other hand, the “Anxiety & Pain” cluster shows higher likelihood for the most frequent hospital admissions and A&E arrivals.

Comparing the “Pain” and “Stress & Pain” clusters, despite substantial overlap in over-represented indicators, the latter group comprises a higher proportion of females. Notably, the overrepresented levels of inpatient and outpatient admissions are lower in the “Stress & Pain” cluster, while both the WIMD and the condition accrual gap are higher. Relative to “Anxiety & Pain” and “Depression & Pain” clusters, individuals in “Stress & Pain” group exhibit less severe interactions with the health care system, fewer LTCs, and reside in less deprived areas.

Some burden indicators appear to be closely linked within the identified clusters. For example, the number of long-term conditions (LTCs), the number of medications, and the number of GP interactions tend to follow the same trend—when a higher level of one is over-represented in a cluster, a higher level of the others also tends to be over-represented.

Less frequently recorded indicators may help to shape different burden profiles or clusters. For instance, in clusters where separation/divorce is over-represented, depression is consistently also over-represented. Over-representation of work change and pain also frequently co-occur in clusters. Mobility problems co-occur with these two burdens in all their instances except for cluster 9.

To our knowledge ours is the first study to cluster individuals with MLTC on the basis of the pattern of burden they experience. This patient-centred approach is novel and highlights the relationship between health inequalities, quality of life and socioeconomic deprivation.

However, the patient experience of burden is only partially captured in routine EHR data. Fraser et al [29] discuss limitations and bias of EHR data in capturing patient-focused burden concepts. We highlight that our binary burden indicators, derived from clinical code lists, do not directly capture information about the patient, but are mediated through the coding practice of their healthcare providers. Thus, if, for example, a cluster defined by a coding of stress in the absence of pain has a higher prevalence of patients from the least deprived areas, this may reveal something about the experience of higher-income patients, but it may also reflect patterns in how GPs tend to code information about higher-income patients. Similarly, prescribing and referral patterns may vary by patient demographic characteristics, by GP characteristics, or as a result of the interaction between the two. For example, some patient groups may be more likely to successfully advocate for themselves to receive or reduce medications, or younger GPs may have different prescribing practices from older ones. At present we cannot reliably untangle the contributions from patient-related and healthcare-provider-related information that are reflected in the EHR.

Furthermore, patient burden is only captured in EHR data when patients interact with healthcare systems. Individuals who interact with the system less frequently may be less likely to have burden indicators recorded. Thus there may be a bias towards under-recording of burden in demographic groups that access healthcare less frequently. For example, it is well-documented that men are less likely to use primary care services [51, 52], and this could be one reason why males are over-represented in the

“Low burden” cluster and females are over-represented in nearly all the others. In addition, if the recording of any burden indicators is itself biased according to these same patient characteristics, this could either counteract or compound differences arising from the different frequency of healthcare interactions. For example, if a particular burden indicator is more likely to be recorded for females than males (e.g. anxiety [53]), then this would compound any difference arising from men being less likely to consult. This potential for ‘hidden’ burden in large ostensibly healthier groups is important to take into account when planning healthcare interventions to support vulnerable and higher-burden groups.

Our analysis can be repeated with more comprehensive, tailored and reliable indicators of patient burden as they become available and routinely recorded. It can also be adapted to focus on particular aspects of the patient experience by curating a bespoke set of indicators, or on particular subpopulations of patients, for example those with particular demographic characteristics, or living in a specific region, or with particular long-term conditions.

A MELD-B Delphi study on the importance of aspects of patient workload associated with MLTC, and the frequency of coding or ability to calculate related concepts in EHR, has recently been completed. It reached consensus with people living with MLTC, carers, clinicians and expert MLTC-researchers on a set of indicators of patient burden that can be used in analyses such as ours.

Profiling patient burden, especially if coupled with focused and careful collection of data that accurately captures the patient experience, opens the way to identifying how health services might best be delivered to ease patients’ experience of managing long-term conditions and improve quality of life.

## Data Availability

Data may be obtained from a third party and are not publicly available. The data used in this study are available in the SAIL Databank at Swansea University, Swansea, UK. Applications to access data via SAIL can be made following their established process https://saildatabank.com/data/apply-to-work-with-the-data/.

## Acknowledgments

We would like to acknowledge all other members of the MELD-B Consortium and additional PPI contributors. We gratefully acknowledge the patients and data providers who make anonymized data available for research.

## Footnotes

1 Currently, GP visits cannot be distinguished from other interactions.

